# Feasibility of integrating a point of care triage test into routine antiretroviral therapy monitoring in Mozambique: a qualitative evaluation

**DOI:** 10.64898/2026.05.19.26353112

**Authors:** Hanlie Myburgh, Anna Saura-Lázaro, Erika van den Bogaart, Denise Naniche, Dulce Adolfo Bila, Madina Ficher-Cunhete, Arminda Ubisse, José Pembelane, Paula Vaz, Rene Paulussen, Lario Viljoen

## Abstract

**Introduction:** Viral load monitoring is central to assessing antiretroviral therapy (ART) effectiveness, yet timely access remains challenging in resource-constrained settings. Point-of-care (POC) triage tests may improve ART monitoring efficiency by identifying clients requiring confirmatory viral load testing while reducing unnecessary testing among those likely to be virally suppressed. We explored perceptions of integrating a POC triage test that measures interferon-gamma-induced protein 10 (IP-10) – a chemokine strongly correlated with HIV viral load – into routine ART monitoring among people living with HIV (PLHIV) on ART, healthcare providers, and HIV programme stakeholders.

**Methods:** This qualitative study was nested within a clinical evaluation of the IP-10 POC triage test in two primary healthcare facilities in Maputo Province, Mozambique (2023–2024). We conducted three rounds of interviews with PLHIV on ART who underwent IP-10 testing, and one-off interviews with healthcare providers and HIV programme stakeholders across different health system levels. PLHIV were purposively sampled to capture diverse IP-10 and viral load outcomes. Interviews explored experiences of ART monitoring, perceptions of the IP-10 POC test, and implementation considerations. Data were analysed thematically using an inductive-deductive approach.

**Results:** Routine viral load monitoring was widely valued and understood as essential for treatment adherence and effectiveness, but participants described barriers including laboratory delays, access challenges, and health system constraints. The IP-10 POC triage test was broadly acceptable; same-day results were perceived to reduce anxiety, support adherence, and enable timely clinical decision-making. Providers and stakeholders emphasised its potential to improve monitoring efficiency by prioritising clients who require confirmatory viral load testing and adherence support. Concerns were raised regarding test accuracy and the need to maintain confirmatory viral load testing, underscoring the importance of clear communication and client education. Successful implementation would require training, workflow integration, and quality assurance.

**Conclusions:** An IP-10 POC triage test could strengthen ART monitoring by enabling same-day identification of clients requiring confirmatory viral load testing and targeted adherence support. By reducing unnecessary viral load testing for virally suppressed clients, it may contribute to more efficient monitoring and support differentiated care approaches. Careful integration into existing ART monitoring algorithms will be critical to maximise impact.

## 1. Introduction

Effective monitoring of antiretroviral therapy (ART) is essential to ensure treatment success and long-term viral suppression among people living with HIV (PLHIV). Viral load measurement – the quantification of HIV viral RNA in blood – remains the gold standard for assessing ART effectiveness and identifying treatment failure and suboptimal adherence (1–3). However, implementing routine ART monitoring through viral load has become increasingly complex following the World Health Organization’s (WHO) 2015 recommendation to adopt a universal ‘treat all’ approach (1, 4). This policy shift expanded ART eligibility to all PLHIV regardless of clinical stage, increasing global ART coverage from 46% in 2015 to 77% in 2024 (5, 6). As intended, individuals are initiating ART earlier in disease progression, improving health outcomes and longevity. However, approximately 30.7 million of the 39.9 million PLHIV globally now require ongoing ART monitoring, including annual viral load measurement (5, 7).

Sub-Saharan Africa, which carries the largest share of the global HIV burden, faces particular challenges in sustaining high-quality ART monitoring (2). Standard viral load testing relies on venous blood collection and centralised laboratory analysis, which are resource-intensive and dependent on efficient specimen referral systems. Many countries in the region continue to face limited laboratory capacity, shortages of trained personnel, and logistical constraints affecting specimen transport and result return. Turnaround times often range from days to weeks, delaying clinical decision-making and undermining client engagement. For many clients, particularly in rural or resource-limited areas, long travel distances and out-of-pocket costs further impede timely result collection and continuity of care (1, 3, 8, 9).

These realities highlight the need for innovative approaches that can rapidly and affordably identify individuals at risk of treatment failure and prioritise them for confirmatory viral load testing (2, 10). A reliable point-of-care (POC) or near-POC triage test could help streamline ART monitoring by distinguishing PLHIV on ART likely to be virally suppressed from those requiring further investigation (11). Developing and validating feasible triage tools represents an important step toward strengthening differentiated care models and sustaining treatment success in high-burden, resource-limited settings.

In this paper, we present qualitative findings from the VIP2 study, which assessed the specificity, acceptability, and feasibility of a POC test to triage PLHIV on ART for viral load testing during routine ART monitoring in Mozambique. While performance characteristics of triage tools are increasingly studied, less is known about how such tools are understood, trusted, and integrated within existing care practices.

## 2. Methods

### 2.1. Study design and setting

A descriptive, exploratory study to assess the acceptability and feasibility of a POC test to triage PLHIV on ART requiring viral load testing using semi-structured interviews with end users (healthcare providers and PLHIV on ART) and HIV programme stakeholders between 2023 and 2024. Two primary healthcare facilities in the Magude and Matola districts of Maputo province, southern Mozambique, were included – one rural and one urban – respectively. With 15.4%, the province had the third highest HIV prevalence in the country in 2021 (12). In 2021, the included health facilities reported viral load testing coverage of between 80-85% and a viral load suppression rate of 78-81% (13).

### 2.2. The VIP2 study

This qualitative assessment was nested in the clinical performance evaluation of a POC lateral flow assay measuring interferon gamma-induced protein 10 (IP-10), a chemokine strongly correlated with HIV viral load (the VIP2 study). The IP-10 POC triage test uses finger-prick blood to assess IP-10 concentration within 20 minutes and does not require specialised equipment or personnel, making it suitable for decentralised use. Previous evaluations of the IP-10 test demonstrated high sensitivity, moderate specificity, and cost-effectiveness (11).

This qualitative study assessed the acceptability and feasibility of integrating the IP-10 POC triage test into Mozambique’s two-step ART monitoring algorithm. Mozambican guidelines align with WHO recommendations for viral load testing at six and twelve months after ART initiation, and annually thereafter for individuals stable on ART (Figure 1). If the viral load is unsuppressed (>1000 copies/mL), three monthly enhanced adherence counselling (EAC) sessions are recommended, followed by repeat viral load testing (14).

**Figure 1.**
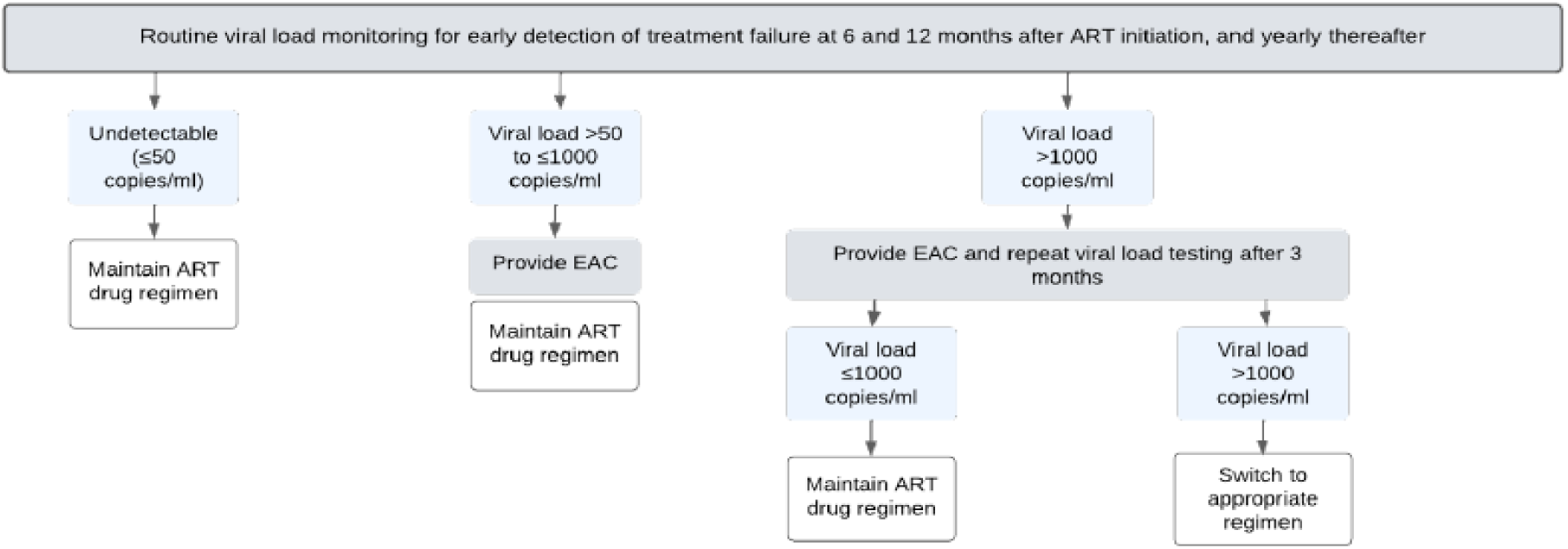
Mozambique’s ART monitoring algorithm, aligned with current WHO recommendations. ART = antiretroviral therapy; EAC = Enhanced Adherence Counselling

As shown in Figure 2, integrating the IP-10 POC triage test into the ART monitoring algorithm would allow PLHIV with negative IP-10 results to forgo further viral load testing, reserving viral load tests for those screening IP-10 positive. This approach could enable same-day ART monitoring for most PLHIV on ART while reducing the number of viral load tests and follow-up visits among virally suppressed (<1000 copies/mL) clients, potentially improving retention in care and reducing health system costs.

**Figure 2.**
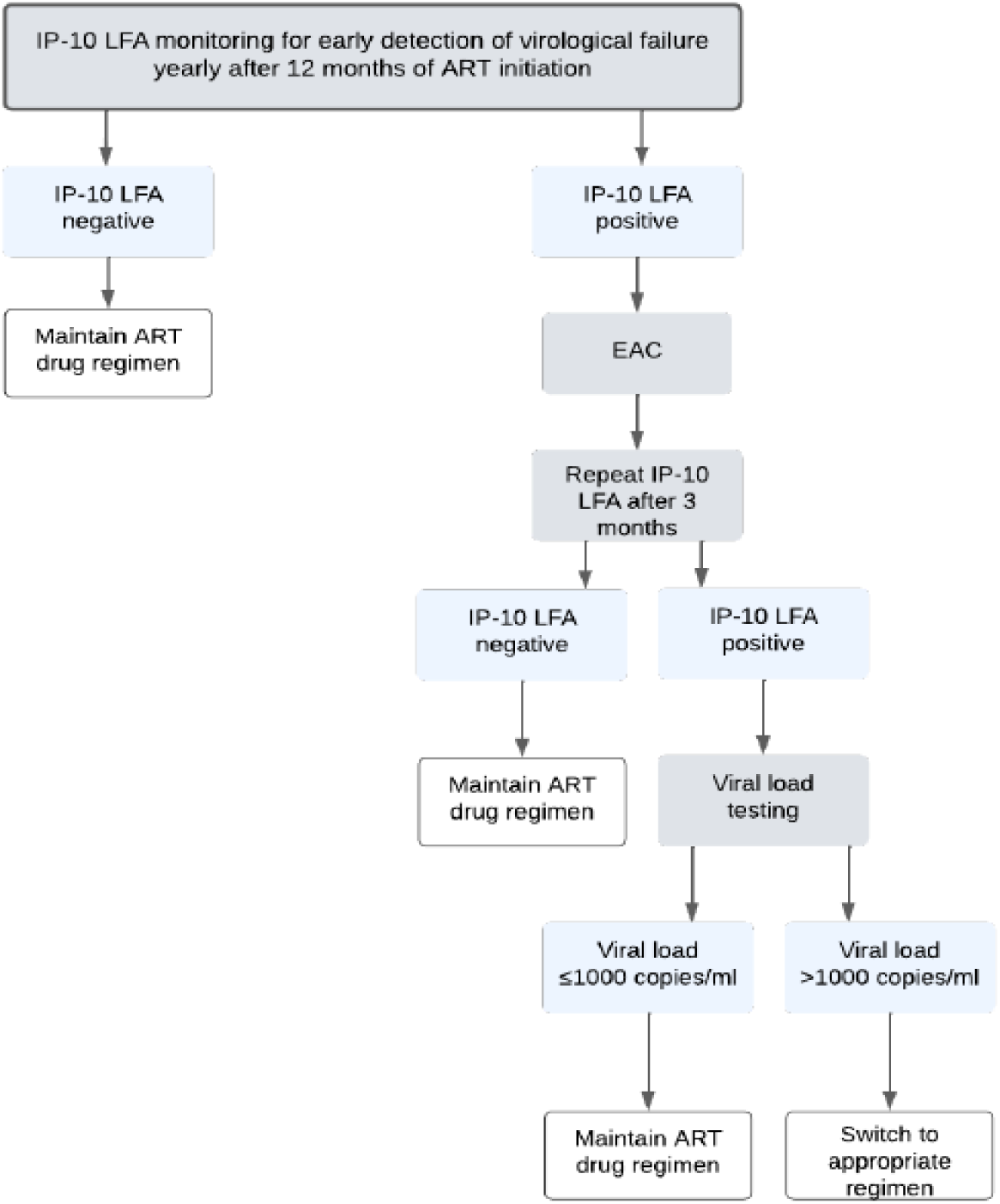
Proposed integration of the IP-10 POC triage test in Mozambique’s routine ART monitoring algorithm. IP-10 LFA = IP=10 lateral flow assay; ART = antiretroviral therapy; EAC = Enhanced Adherence Counselling

### 2.3. Sampling, recruitment, and data collection

We conducted interviews with three participant groups: PLHIV on ART, healthcare providers, and HIV programme stakeholders. Two research assistants experienced in qualitative methods and the Mozambican health and social context conducted interviews with PLHIV on ART and healthcare providers. A study researcher with extensive experience in supporting the implementation of the Mozambican HIV programme conducted stakeholder interviews. All participated in a week-long training workshop before data collection.

#### 2.3.1. PLHIV on ART

Data collection with PLHIV on ART was embedded within the clinical performance assessment across three rounds of interviews (Figure 3). We used an adaptive, open-cohort design to capture diverse experiences across IP-10 and viral load outcomes and minimise attrition. Participants were sampled from both healthcare facilities. Recruitment in the clinical performance study included clients attending stable and specialized risk clinics to enrich enrolment of PLHIV likely experiencing treatment failure or suboptimal adherence. Rounds 1 and 3 coincided with routine ART monitoring visits. Clinical management followed national guidelines, including EAC and 90-day follow-up visits for PLHIV on ART with unsuppressed viral load. During Round 1 (study visit 1), participants received a finger-prick IP-10 POC triage test and routine venous blood draw for viral load testing, conducted by a study nurse (15). An interim analysis identified the optimal IP-10 threshold for the qualitative study, enabling participants to receive an IP-10 positive or negative result.

**Figure 3.**
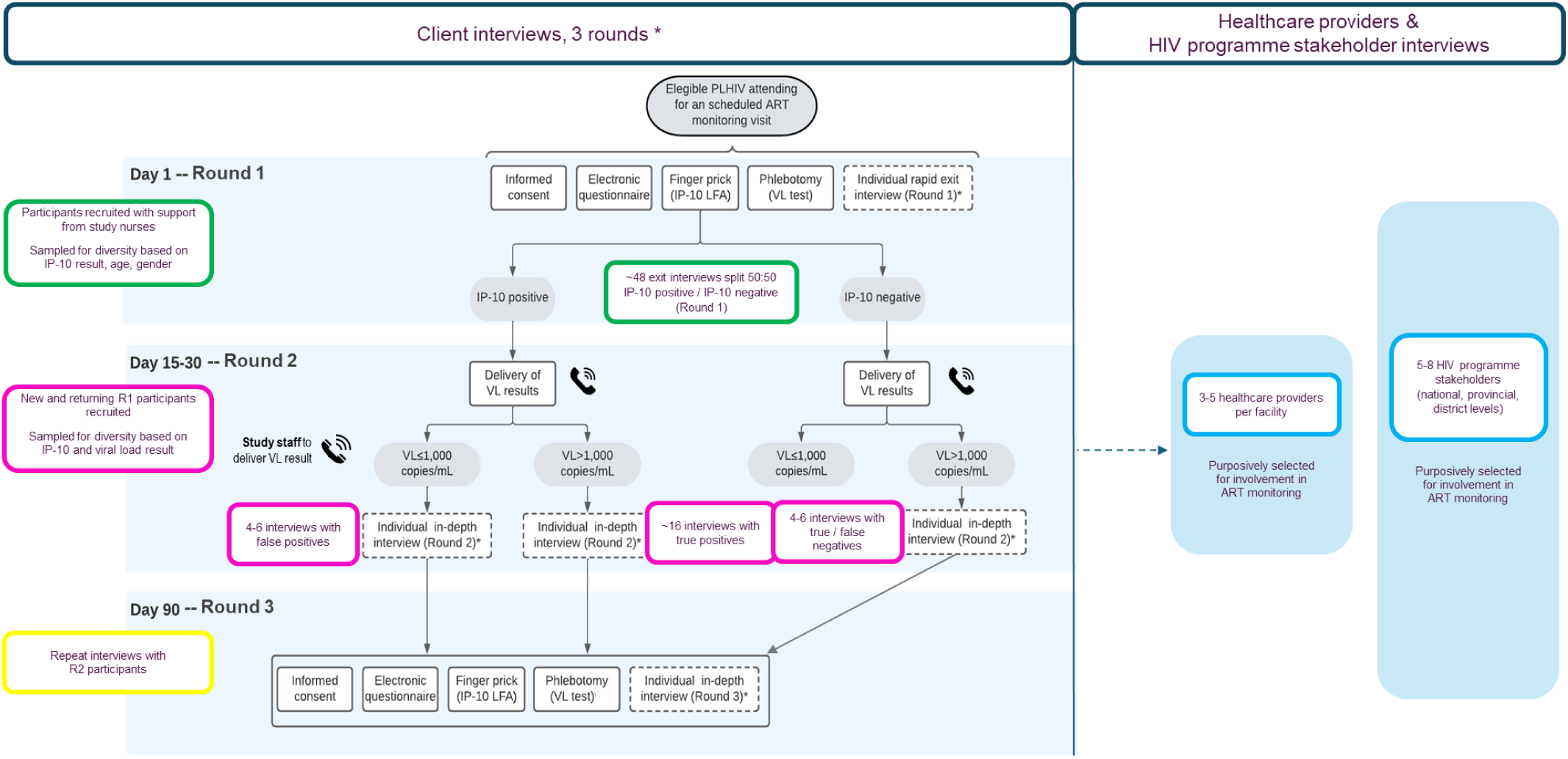
Sampling, recruitment, and data collection alongside the IP-10 POC triage test’s clinical performance assessment. VL = Viral Load; PLHIV = people living with HIV; IP-10 LFA = IP-10 Lateral Flow Assay; ART = antiretroviral therapy

We aimed to recruit 48 participants in Round 1 for rapid exit interviews exploring immediate perceptions of the IP-10 POC triage test, purposively sampling by age (18-24; 25-40; >40 years), gender, and IP-10 result.

Following receipt of viral load test results (15–30 days later), approximately 28 participants were recruited for in-depth interviews (Round 2) exploring HIV treatment journeys, understandings of ART monitoring, and experiences of the IP-10 POC test. To capture diverse perspectives, we included participants with true positive (IP-10 positive, viral load >1000 copies/mL), false positive (IP-10 positive, viral load ≤1000 copies/mL), false negative (IP-10 negative, viral load >1000 copies/mL), and true negative (IP-10 negative, viral load ≤1000 copies/mL). Both Round 1 and newly recruited participants could participate if they had received both IP-10 and viral load results. In Round 3 (study visit 3), we re-engaged Round 2 participants to explore ART adherence, EAC experiences (where relevant), and evolving perceptions of ART monitoring and the IP-10 POC test.

All interviews were conducted in private spaces at the healthcare facilities, using study-specific topic guides tailored to each round. Interviews were conducted in participants’ preferred language, audio recorded, and ranged from 10-20 minutes for exit interviews and 30-90 minutes for in-depth interviews.

#### 2.3.2. Healthcare providers

We purposively sampled 3-5 healthcare providers per facility, including clinicians, psychologists, counsellors, and study nurses involved in ART monitoring and patient support. Recruitment occurred in 2024 midway through the clinical performance study, when providers were already familiar with the VIP2 study. Providers participated in one-off private interviews covering participant background, community context, facility-level ART monitoring, and perceptions of the IP-10 POC triage test. Interviews were conducted in participants’ preferred language, audio recorded, and lasted 30-90 minutes.

#### 2.3.3. HIV programme stakeholders

We aimed to interview 5-8 HIV programme stakeholders from district, provincial, and national levels, purposively selecting for involvement in ART monitoring. Interviews explored strengths and limitations of the current ART monitoring strategy, perceptions of integrating the IP-10 POC triage test into the national algorithm, and implementation considerations. Participants were recruited via e-mail or telephone. Interviews were conducted in person or online, in participants’ preferred language, audio recorded, and lasted between 45-60 minutes.

### 2.4. Data analysis

Research assistants produced detailed case descriptions of interview recordings in the original language using structured templates tailored to each participant group. Templates captured participant background, summaries of key discussion topics supported by excerpts and observations, and researcher reflections on the interview’s contribution to study objectives. The summary approach, previously used across multiple settings, is less time-consuming than full transcription while maintaining comparable quality (16). Case descriptions were translated into English using online tools and quality checked by a local language speaker with HIV programme experience. These summaries served as the primary analytical material.

We used a combined deductive-inductive coding approach in ATLAS.ti. Initial coding used deductively defined labels capturing experiences of health services, perceptions of the IP-POC test, and implementation considerations. Codes were then inductively refined through an analysis workshop involving the field team and study authors. Preliminary findings were discussed in a reflexive team workshop led by research assistants, and analysis was further refined through iterative writing and review across the wider study team.

### 2.5. Ethics approval and consent to participate

Ethics approval for this project was granted through the bioethics committees for health of the Mozambique National Health Institute (Comité Nacional de Bioética para a Saúde – CNBS) and Autoridade Reguladora de Medicamento, IP (ANARME). An informed consent process for the qualitative component, in addition to that of the main clinical study, was followed with all participants prior to participating in an interview, after which participants were asked to complete a written informed consent form. The informed consent process and written informed consent form were available in the local language/s.

## 3. Results

We present an overview of participants across the three sampling categories, outline the broader HIV care context as described by participants, and examine perceptions of the IP-10 POC triage test and its potential integration into the national ART monitoring algorithm. We also highlight key considerations for implementing a POC viral load triage test during routine ART monitoring. Interview excerpts illustrate perspectives relevant to the study objectives, rather than prevalence.

### 3.1. Participant overview

#### 3.1.1. PLHIV on ART

A total of 43 PLHIV on ART participated in exit interviews in Round 1 across the two healthcare facilities (Figure 4). In Round 2, 29 interviews were conducted, including 13 participants who had also taken part Round 1. Thirteen (n=13) interviews were completed in Round 3, all with participants previously interviewed in Round 2. Recruitment in Round 3 was lower than anticipated due to attrition and condensed study timelines.

**Figure 4.**
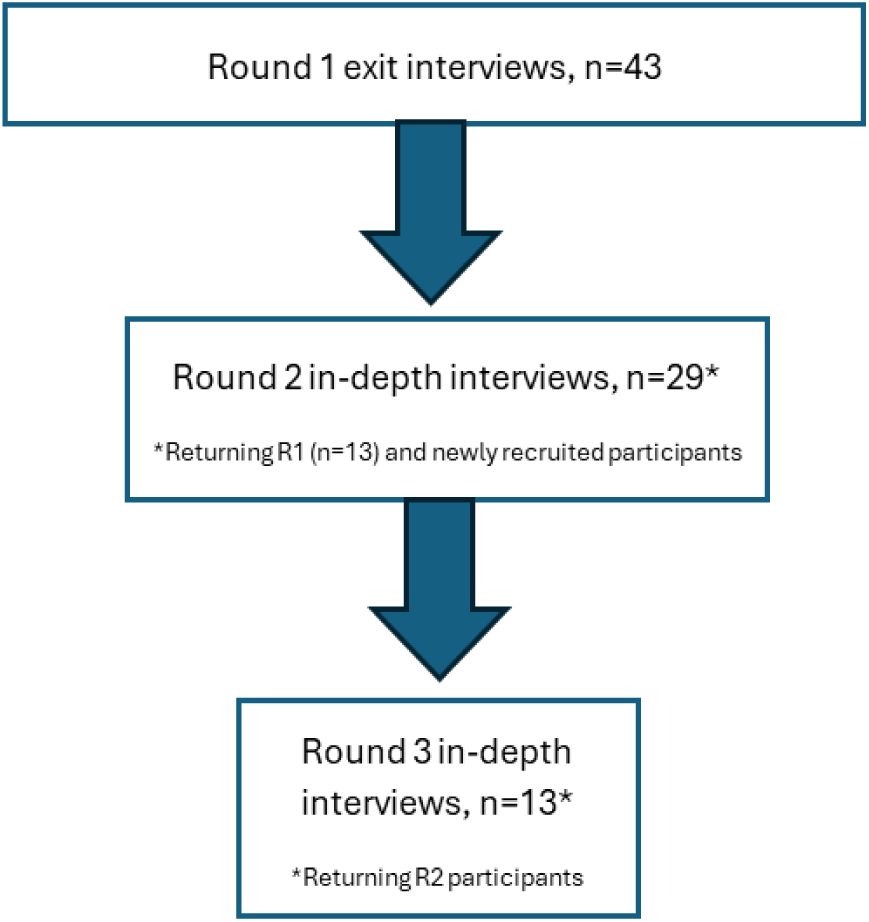
Number of participants enrolled per data collection round across the two healthcare facilities.

Table 1 details participant demographics, time on ART, reported adherence, and IP-10 POC test results for Round 1 participants; these findings are reported elsewhere (15). More women (n=28, 65%) than men participated, and most participants were aged 26–49 years (n=31, 72%), reflecting the overall client populations at both facilities. Although Round 1 sampling aimed for equal numbers of participants with positive and negative IP-10 POC test results, more participants received positive results due to the test’s moderate specificity (15). These proportions were also similar to those in the overall VIP2 study population.

**Table 1.**
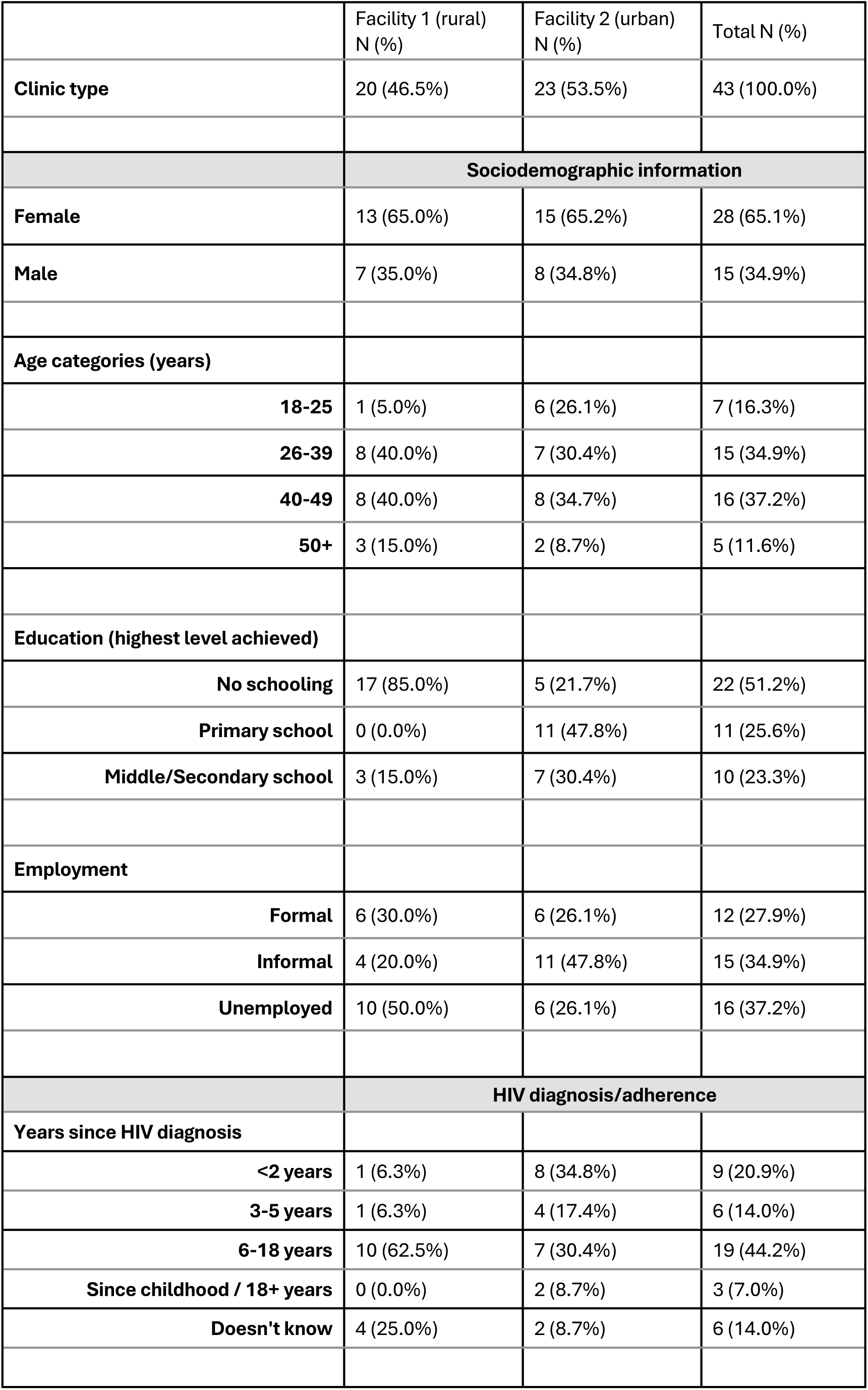
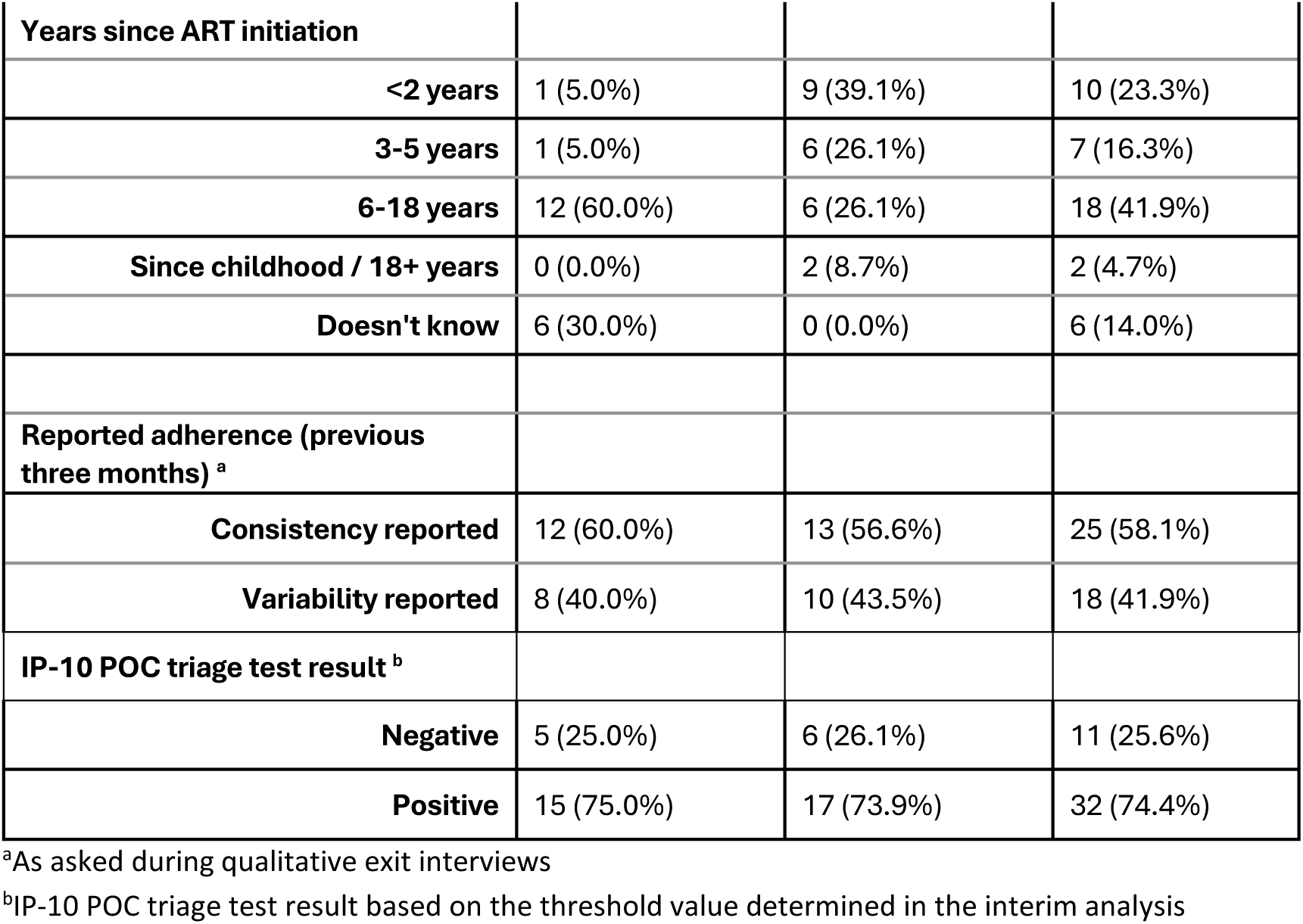
Profile of sampled PLHIV on ART interviewed in exit interviews referred to as Round 1, according to the healthcare facility.

|  | Facility 1 (rural)<br>N (%) | Facility 2 (urban)<br>N (%) | Total N (%) |
| --- | --- | --- | --- |
| **Clinic type** | 20 (46.5%) | 23 (53.5%) | 43 (100.0%) |
|  | **Sociodemographic information** |  |  |
| **Female** | 13 (65.0%) | 15 (65.2%) | 28 (65.1%) |
| **Male** | 7 (35.0%) | 8 (34.8%) | 15 (34.9%) |
| **Age categories (years)** |  |  |  |
| **18-25** | 1 (5.0%) | 6 (26.1%) | 7 (16.3%) |
| **26-39** | 8 (40.0%) | 7 (30.4%) | 15 (34.9%) |
| **40-49** | 8 (40.0%) | 8 (34.7%) | 16 (37.2%) |
| **50+** | 3 (15.0%) | 2 (8.7%) | 5 (11.6%) |
| **Education (highest level achieved)** |  |  |  |
| **No schooling** | 17 (85.0%) | 5 (21.7%) | 22 (51.2%) |
| **Primary school** | 0 (0.0%) | 11 (47.8%) | 11 (25.6%) |
| **Middle/Secondary school** | 3 (15.0%) | 7 (30.4%) | 10 (23.3%) |
| **Employment** |  |  |  |
| **Formal** | 6 (30.0%) | 6 (26.1%) | 12 (27.9%) |
| **Informal** | 4 (20.0%) | 11 (47.8%) | 15 (34.9%) |
| **Unemployed** | 10 (50.0%) | 6 (26.1%) | 16 (37.2%) |
|  | **HIV diagnosis/adherence** |  |  |
| **Years since HIV diagnosis** |  |  |  |
| **<2 years** | 1 (6.3%) | 8 (34.8%) | 9 (20.9%) |
| **3-5 years** | 1 (6.3%) | 4 (17.4%) | 6 (14.0%) |
| **6-18 years** | 10 (62.5%) | 7 (30.4%) | 19 (44.2%) |
| **Since childhood / 18+ years** | 0 (0.0%) | 2 (8.7%) | 3 (7.0%) |
| **Doesn't know** | 4 (25.0%) | 2 (8.7%) | 6 (14.0%) |

| Years since ART initiation |  |  |  |
| --- | --- | --- | --- |
| <2 years | 1 (5.0%) | 9 (39.1%) | 10 (23.3%) |
| 3-5 years | 1 (5.0%) | 6 (26.1%) | 7 (16.3%) |
| 6-18 years | 12 (60.0%) | 6 (26.1%) | 18 (41.9%) |
| Since childhood / 18+ years | 0 (0.0%) | 2 (8.7%) | 2 (4.7%) |
| Doesn't know | 6 (30.0%) | 0 (0.0%) | 6 (14.0%) |
| Reported adherence (previous three months) <sup>a</sup> |  |  |  |
| Consistency reported | 12 (60.0%) | 13 (56.6%) | 25 (58.1%) |
| Variability reported | 8 (40.0%) | 10 (43.5%) | 18 (41.9%) |
| IP-10 POC triage test result <sup>b</sup> |  |  |  |
| Negative | 5 (25.0%) | 6 (26.1%) | 11 (25.6%) |
| Positive | 15 (75.0%) | 17 (73.9%) | 32 (74.4%) |

<sup>a</sup>As asked during qualitative exit interviews
<sup>b</sup>IP-10 POC triage test result based on the threshold value determined in the interim analysis

Round 2 participants (n=29) had a similar gender and age distribution to Round 1. To capture perspectives across different IP-10 and viral load results (Table 2), the sample included participants with true positive (n=15), true negative (n=3), false positive (n=8), and false negative (n=3) IP-10 POC test results. The Round 3 sample showed a similar distribution of IP-10 and viral load results (Table 2).

**Table 2.** IP-10 and HIV viral load result distribution of sampled PLHIV on ART interviewed in Round 2 and Round 3.

|  | Round 2<br>N=29 | Round 3<br>N=13 |
| --- | --- | --- |
| Women | 20 (69.0%) | 10 (77.0%) |
| Mean age (years) | 38 | 38 |
| True positive<br>(+) IP-10 test, VL >1,000 copies/mL | 15 (51.7%) | 7 (53.9%) |
| True negative<br>(-) IP-10 test, VL ≤1,000 copies/mL | 3 (10.3%) | 2 (15.4%) |
| False positive<br>(+) IP-10 test, VL ≤1,000 copies/mL | 8 (27.6%) | 3 (23.1%) |
| False negative<br>(-) IP-10 test, VL >1,000 copies/mL | 3 (10.4%) | 1 (0.1%) |

\*VL = viral load

#### 3.1.2. Healthcare providers

We interviewed ten healthcare providers across the two healthcare facilities, including a medical officer, an ART nurse, a psychologist/counsellor, a laboratory technician, and a study nurse at each facility. Eighty per cent (n=8) were women, with a mean age of 35 years.

#### 3.1.3. HIV programme stakeholders

We interviewed six stakeholders involved in the HIV programme at district (n=2), provincial (n=1), and national level (n=3), variably representing the Sexually Transmitted Infections (STIs) & HIV/AIDS Control Programme and Diagnostics and Laboratory.

### 3.2. HIV care context: health service delivery and routine ART monitoring

#### 3.2.1. PLHIV on ART

Participants described largely positive experiences of HIV care, emphasising respectful communication, clear explanations, and ongoing support. Nurses were trusted sources of information, helping patients understand HIV and treatment adherence. Most participants demonstrated a basic understanding of HIV and ART, describing the virus as ‘bugs’ and ART as reducing the number of ‘bugs’ in the blood. Healthcare providers explained that such concepts were used to support treatment ownership among clients with low literacy. As one woman noted: ‘*It was the nurses who explained [HIV] to me. They speak well. When you have any doubts, you can ask*’ (woman, 18-25 yrs, Facility 1, rural).

HIV viral load testing was understood as central to treatment success, and participants reported that results were routinely shared and explained. Providers often linked missed doses or alcohol use to viral rebound. As a client reflected, ‘*If you don’t medicate properly, if you consume alcohol, your viral load will rise, and those bugs will be in greater quantity than the soldiers [CD4 cells]’* (man, 40-49 yrs, Facility 1, rural). In the rural facility, some participants expressed mistrust about how blood samples were used, including concerns that blood might be sold.

Many participants described feeling cared for beyond routine consultations, including follow-up calls about adherence, nutrition, and well-being. Disclosure to trusted family members was common, and relatives often assisted with medication collection. Adherence support services were experienced ambivalently: while counseling groups were valued by some, others viewed referral to counsellors as a form of ‘punishment’ for missed doses that increased the number of clinic visits. Overall, ART monitoring was understood as both a clinical and moral practice, with viral load results functioning not only as biomedical indicators but also as markers of responsibility, adherence, and being ‘well’ in the eyes of providers, as a participant explained: ‘*[The healthcare providers] tell us if we are medicating properly when the results of the blood samples come out…They guide me to follow good paths…If you kill yourself, that…has to do with you’* (woman, 26-39 yrs, Facility 1, rural).

#### 3.2.2. Healthcare providers

Healthcare providers described multiple challenges undermining treatment adherence, retention, and monitoring. Distance from facilities and travel costs limited attendance, while low HIV literacy and persistent myths about HIV and ART remained common. At the rural facility, providers described highly mobile seasonal farm worker communities, making follow-up and retention difficult. Harmful alcohol and drug use further complicated care delivery and adherence. Providers explained that, in these constrained contexts, clients often made pragmatic decisions about treatment:

> ‘*Sometimes, patients don’t have medication, but they stay home [because they don’t have means to travel to the facility],*’ (Counsellor, Facility 1, rural).
>
> *‘[This client who did not return to the facility for his viral load result] will continue to take [ART], but without knowing his condition,’* (ART nurse, Facility 1, rural).

Annual viral load testing through routine blood draws was the primary method for monitoring adherence and treatment response. Providers described differentiated service delivery models, including community medication distribution for stable clients and intensified follow-up for those with elevated viral loads through EAC, repeat viral load testing, and telephonic or home visits.

Providers also highlighted systemic and resource-related constraints affecting HIV service delivery and ART monitoring. Understaffed, under-resourced facilities contributed to long waiting times, while power interruptions (particularly at the rural site) and limited laboratory infrastructure delayed viral load testing. Laboratory challenges, including misplaced specimens and delayed results, often required additional facility visits. Some providers expressed embarrassment when blood tests had to be repeated because of service failures, noting that this undermined client trust and fuelled suspicions about the use of their blood samples.

#### 3.2.3. HIV programme stakeholders

HIV programme stakeholders across health system levels described the national ART monitoring strategy as aligned with global guidance, including U=U (undetectable equals untransmissible), and supportive of achieving undetectable viral load among PLHIV on ART. They emphasised the value of routine viral load monitoring for assessing treatment response and disease progression, alongside interventions such as EAC to support viral suppression.

Despite describing the national strategy as robust, stakeholders identified barriers to effective implementation, including limited laboratory capacity and human resources, high staff turnover, and missed opportunities to conduct viral load tests according to national guidelines. National-level stakeholders particularly highlighted poor adherence to the ART monitoring algorithm, noting persistent delays in providers ordering viral load tests: ‘*We have observed this in the monthly reports and in the quality improvement data, which show the same fragility in the request for viral load’* (Stakeholder, national).

### 3.3. Acceptability of an IP-10 POC test to triage viral load testing during routine ART monitoring

In the context of the challenges related to HIV services delivery and access, all three participant groups welcomed the idea of a POC test to triage viral load testing as part of routine ART monitoring.

#### 3.3.1. PLHIV on ART

Immediate acceptability, assessed through Round 1 exit interviews, showed strong acceptance among PLHIV on ART. Participants welcomed the IP-10 POC triage test as less invasive, less painful, and a more immediate indicator of health during clinic visits. Positive perceptions continued in in-depth interviews, where many understood the test as a rapid indicator of adherence and overall health. Receiving results on the same day was described as a major advantage, reducing anxiety about treatment effectiveness and eliminating the usual delay associated with routine viral load testing.

Participants demonstrated a strong desire to understand their health status and openness to tools that could support this. Many reported that immediate results motivated adherence by either confirming good practice or signalling the need for improvement: ‘*The finger test is good because the result does not take long. If you are wrong, you can immediately correct what is wrong,’* (man, 40-49 yrs, Facility 1, rural). Participants also highlighted the test as substantially less painful and invasive than venous blood draws, often comparing it to familiar rapid tests. As one woman noted, ‘*it’s just a prick, as if I were taking a malaria test,’* (woman, 26-39 yrs, Facility 1, rural). Another likened it to HIV testing: *‘it’s the same as the HIV test…you do it today and get [the result] today,*’ (woman, 26-39 yrs, Facility 2, urban).

Despite generally positive perceptions, participants expressed concerns about the test’s accuracy, reflecting the limitation that IP-10 is not specific to viral load. False positive results (a positive IP-10 POC triage test followed by an undetectable viral load result (<50 copies/mL)) sometimes caused distress, with participants interpreting the result as evidence of worsening health or poor HIV control. Some were also uncomfortable with the idea of replacing the ‘formal’ viral load blood draw, which they trusted more because of laboratory processing and more rigorous specimen collection. As one participant explained, ‘*You can get the result as quickly as possible, and you will know how to control yourself, but it is always better to [draw blood from] the arm to be sure of the result’* (man, 40-49 yrs, Facility 2, urban). Another added: ‘*The fact that it is fast does not do much good because it is better to have time for them [laboratory and clinical staff] to investigate the correct and true result,’* (woman, 50+ yrs, Facility 2, urban).

Participants also described high levels of trust in healthcare providers, reinforcing providers’ role as key mediators of confidence in new tools and services. As one participant explained: ‘*I would take the test again… I simply fulfil everything that is said to me at the hospital. If they send me somewhere, I go without question,’* (woman, 26-39 yrs, Facility 1, rural).

#### 3.3.2. Healthcare providers

Healthcare providers identified the rapid turnaround time of the IP-10 POC test and its potential to facilitate timely clinical decision-making as key advantages. They noted that same-day results could help address some challenges associated with care delivery in surrounding communities:

> *‘It’s very good, because we can intervene immediately. I think it’s good just to know [the result] in 20 minutes,*’ (Psychologist, Facility 2, urban).
>
> *‘It would be good because…each patient will already return home with their result,’* (Medical officer, Facility 1, rural).

Although providers received information about the IP-10 POC triage test and the proposed ART monitoring algorithm, many had a limited understanding of the test’s mechanism or the clinical implications of its results. Nevertheless, they recognised its potential to improve ART monitoring efficiency by reducing venous blood draws. Only laboratory technicians demonstrated a clear understanding of IP-10 and its relationship to elevated viral load, shaping more nuanced perspectives on the test’s accuracy and limitations. As one technician explained:

*‘It is a non-accurate test, but it is accurate [in what it measures, i.e., IP-10] …When done, the quick results already tell you that something is not right with a patient… It would serve as a warning to [clinicians and] patients without having to wait for the results from [the laboratory],’* (Laboratory technician, Facility 1, rural).

#### 3.3.3. HIV programme stakeholders

Stakeholders generally viewed the IP-10 POC triage test favourably, particularly for its potential to address limitations in the current ART monitoring strategy. Perceived advantages included reducing missed opportunities to assess adherence during facility visits, enabling more timely support interventions, and decreasing logistical complexity and costs by screening out clients who may not require laboratory-based viral load testing. As one provincial stakeholder noted:

> *‘Instead of worrying about 100 patients, I’m going to worry about 10…I will already have information for this patient, on the very day that we have this test,’* (Stakeholder, province).

At the same time, stakeholders identified important limitations, including the test’s limited specificity for detecting elevated HIV viral load and its inability to distinguish clinically meaningful viral load thresholds (e.g., 50–1000 copies/mL versus >1000 copies/mL), with implications for clinical decision-making. As another explained:

*‘My biggest concern is that in our current algorithm, the user who has a detectable viral load above 50 copies/mL, I have to work with him because something might be failing… [it’s unclear] with how many [viral load] copies this test comes back positive,’* (Stakeholder, province).

### 3.4. Implementation considerations for an IP-10 POC test to triage viral load testing during routine ART monitoring

The three participant groups shared generally positive views of the IP-10 POC test and its potential to address some limitation of the current ART monitoring strategy based solely on viral load testing. Healthcare providers and stakeholders also proposed implementation considerations involving messaging, training, and impact evaluation to strengthen the acceptability and feasibility of integrating the IP-10 POC triage test into routine ART monitoring. We present implementation considerations for 1) healthcare provider and facility-level implementation, and 2) health system implementation and policy.

#### 3.4.1. At the healthcare provider and facility-level

HIV programme stakeholders and providers highlighted uneven infrastructure across facilities as a major constraint to sustained rollout of new technologies. Reflecting on feasibility, they raised concerns about equipment maintenance, quality monitoring, and performance under high temperatures, drawing on prior experience with diagnostic technologies. As one national stakeholder cautioned:

> *‘The biggest challenge is that in the introduction of any technology, we are not able to maintain the equipment…We have no plans…to monitor quality. So, we will never be able to detect any problem in time to be able to move forward,’* (Stakeholder, national).

Providers also anticipated workflow challenges, including identifying staff responsible for conducting the test, securing appropriate space, and managing client flow during the 20-minute result turnaround time, which requires clients to remain in the consultation room.

Stakeholders also stressed the importance of clear, consistent messaging. The IP-10 POC test should be framed as a triage tool rather than a definitive test to avoid confusion or mistrust, particularly around repeated blood draws. Communication was considered essential for providers, clients, and civil society. Training should emphasise the test’s purpose and limitations to support accurate explanation of results. However, high staff turnover and internal transfers were identified as risks to sustained knowledge and consistent messaging, underscoring the need for ongoing training and system-level support.

#### 3.4.2. For health system implementation and policy

Stakeholders emphasised the need for robust impact evaluation before national adoption, including analysis of performance across populations, cost, clinical outcomes, and effects on quality of care. As one national stakeholder noted: *‘[even an imperfect] test [may] manage to solve a specific problem …the most important thing is to understand how it acts in various situations…which can help design a minimum framework for including in the national health service,’* (Stakeholder, national).

To support adoption, stakeholders highlighted the importance of building acceptability across health system levels to address initial distrust of new technologies. They pointed to existing technical groups as formal platforms for scientific review and policy endorsement, alongside established mechanisms for dissemination to provinces and facilities. Clear positioning was considered critical: the IP-10 POC test should be framed as optimising workflow, strengthening differentiated service delivery, and improving patient comfort and viral load management. Training, they suggested, should be focused and practical, covering what IP-10 measures, its purpose, and guidance on results communication to prevent rumours.

Stakeholders also identified potential value for targeted use in rural or laboratory-constrained settings, among men and key populations with limited access to ART monitoring, and for additional applications such as tuberculosis screening, which also elevates IP-10. Overall, the test was viewed as promising where rapid decision-making and targeted intervention are most needed.

## 4. Discussion

This study examined experiences of routine ART monitoring and the potential role of an IP-10 POC triage test within existing algorithms. Routine monitoring was consistently understood by PLHIV, providers, and programme stakeholders as central to assessing treatment effectiveness and supporting adherence. Viral load testing was highly trusted and reinforced treatment literacy, engagement in care, and motivation to maintain viral suppression. However, persistent structural constraints, including laboratory delays, transport barriers, mobility, and staffing limitations, continued to limit timely and equitable access, reflecting enduring gaps between policy and practice (1, 3, 17–19). Beyond its clinical function, participants’ accounts suggested that ART monitoring also operates as a social and relational process through which treatment engagement and expectations of care are shaped.

Within this context, the IP-10 POC test was understood as a complementary upstream tool rather than a replacement for viral load testing. Same-day results were perceived to enable earlier clinical feedback and more responsive decision-making. Critically, the test supports risk stratification, prioritising confirmatory viral load testing for clients with higher likelihood of viraemia. This signals a shift from standardised monitoring schedules towards risk-stratified approaches that align viral load testing intensity with clinical need, with potential to strengthen differentiated service delivery and optimise constrained laboratory resources (20–23).

Concerns regarding test specificity highlighted both the risks and efficiencies of a triage-based approach. Viral load testing remains the definitive standard for clinical decision-making. However, an appropriately calibrated triage strategy could identify clinically stable clients unlikely to require immediate testing, reducing unnecessary viral load use, costs, and clinic burden. Communication is central to implementation. For PLHIV on ART in our study, the immediacy of triage results enhanced feedback and reinforced engagement in care. Beyond its biomedical function, viral load testing also operated as a relational and moral technology within clinical encounters, shaping how clients understood responsibility, control, and what it means to be ‘well’. These findings resonate with literature showing that viral load functions as a social indicator through which clients are positioned as adherent and responsible (or not) within HIV care (24–26). Within this moral framework, introduction of a triage test may have unintended implications for how care and responsibility are interpreted. The implications of being ‘screened out’ of immediate viral load testing must therefore be framed as an indication of clinical stability rather than withdrawal of care. False positive triage results may cause distress, highlighting the importance of confirmatory testing protocols, robust follow-up systems, and transparent communication of uncertainty. Preferences for venous blood-based testing over finger-prick methods further underscored that trust in diagnostics is shaped by experiential and symbolic factors, not only technical performance. Laboratory-based procedures were often perceived as more reliable, reflecting entrenched hierarchies of diagnostic legitimacy and laboratory authority evidenced elsewhere (27–29). Successful integration of POC tools will therefore depend on both performance and positioning within established practices.

Implementation considerations extended beyond technical performance. Providers and stakeholders emphasised that successful integration would require adequate training, alignment with existing workflows, sufficient infrastructure, and robust quality assurance systems, echoing findings from reviews of POC viral load testing and related implementation research (30). Clear operational guidance on how triage results should trigger confirmatory viral load testing or differentiated follow-up within ART monitoring algorithms will be essential. These findings reinforce broader evidence that diagnostic innovations cannot address gaps in care in isolation but must be embedded within functioning health systems, supported by institutional trust and accompanied by clear operational guidance (31, 32).

Strengths include the inclusion of perspectives across clients, providers, and stakeholders, enabling a multi-level analysis of acceptability and integration. The qualitative design provided in-depth insight into how trust, understanding, and client-provider relationships shape uptake. Limitations include the restricted geographic scope, inclusion of participants already engaged in care, and the research context of test exposure, which may limit transferability. Use of case summaries may have constrained analytic depth, and provider perspectives often reflected anticipated rather than sustained implementation experience.

Overall, the IP-10 triage test has the potential to reshape ART monitoring by enabling more efficient, risk-stratified deployment of viral load testing. As a complementary component within existing algorithms, it may improve timeliness, resource allocation, and client experience. Realisation of this potential will depend on clear protocols, effective communication, and system readiness for implementation at scale.

## Data Availability

All data produced in the present study are available upon reasonable request to the authors

## Authors’ contributions

LV, HM, ASL, EB, and DN designed the research study. DAB, MFC, AU, and JP collected data. All authors analysed the data. HM, LV, ASL, EB, and DN wrote the paper. All authors reviewed and approved the final manuscript.

## Acknowledgements

We would like to acknowledge and thank the Ministry of Health in Mozambique, which approved this research to be conducted, as well as the participating health facilities, healthcare providers, HIV programme stakeholders, and PLHIV on ART at the participating facilities.

## Data availability statement

Anonymised interview transcripts are available on request.

